# Cohort Profile: The Adolescent and Young Adult Tracking Engagement and Management Skills (AYA TEAMS) Longitudinal Cohort of Childhood Cancer Survivors in the United States

**DOI:** 10.64898/2026.02.11.26346092

**Authors:** Sara King-Dowling, Kelsey Woodard, Haley Faust, Sarah Drake, Lina Gov, Dava Szalda, Kemar V. Prussien, Jill P. Ginsberg, Wendy Hobbie, Carole A. Tucker, Lamia P. Barakat, Janet A. Deatrick, Yimei Li, Karen C. Burns, Kathryn Nielsen, Victor Flores, Nithya Ramaswamy, Margaret Jankowski, Bridget O’Hagan, Anneliese Wilkins, David R. Freyer, Ahna Pai, Lisa A. Schwartz

## Abstract

**Purpose:** To describe the rationale, methods, and baseline sample descriptives of the Adolescent and Young Adult Tracking Engagement and Management Skills (AYA TEAMS) cohort. The AYA TEAMS study is a longitudinal observational cohort study that aims to identify determinants and patterns of self-management and engagement in cancer-related long-term follow-up (LTFU) care and validate a novel transition readiness assessment among adolescent and young adult (AYA) survivors of childhood cancer.

**Participants:** AYA survivors of childhood cancer (ages 16-25) and their caregivers were enrolled from 3 large pediatric oncology centers across the United States from 2020-2022 and followed for 2 years (minimum) to 3 years and 3 months (if transferred to adult care). AYA inclusion criteria were: past childhood cancer diagnosis, at least 2 years off-treatment, 5 years since diagnosis, engaged with the participating pediatric health care system within the last 18 months, cognitively able to complete study procedures, and English speaking. AYA completed a comprehensive battery of measures including assessments of self-management and transition readiness at baseline and annually for 2 years. For AYA transferred to adult care, separate measures were administered at the time of transfer (following last pediatric visit) and 15 months post transfer. Caregivers (English or Spanish-speaking) completed a single survey at baseline to capture family functioning, psychosocial risk, and transition readiness. Cancer diagnosis, treatment modalities, treatment-related late effects, and engagement in LTFU care were captured via electronic medical record review. In total, 709 AYA were enrolled and 587 were included in the final cohort [M_age_=19.7 years, 52.5% female, 38.2% from racial and/or ethnic minoritized groups, (REMG)]. The cohort was on average 7.3 years old at the time of diagnosis and 10.5 years off treatment. Half (52.5%) were survivors of leukemia/lymphoma, 38.0% solid tumors, and 9.5% central nervous system tumors. Three hundred and ninety-nine caregivers participated (90% mothers).

**Findings to Date:** Enrolled AYA excluded from the baseline cohort were more likely to be male, from REMG, and/or to enroll without a caregiver. Baseline cohort differences between sites emerged for age, race and ethnicity, socioeconomic status, and treatment modalities and intensity.

**Future Plans:** Data collection was completed in April 2025. Findings from this cohort will elucidate important predictors of self-management and engagement in recommended annual LTFU and inform the design of interventions to reduce disengagement in LTFU.

**Strengths and Limitations:**

- This study is the first known prospective cohort of AYA-only long-term survivors of childhood cancer in the United States recruited from pediatric cancer centers.
- This study achieved high enrollment and retention rates across a medically and demographically diverse sample.
- Informed by multiple theoretical self-management models, this study will be able to examine predictors and transactional relationships of AYA survivor self-management, including engagement in pediatric and adult cancer-related long-term follow-up care.
- Reliance on English-speaking AYA and those currently engaged with the health care system are limitations.

## INTRODUCTION

Over 85% of children diagnosed with cancer become long-term survivors, resulting in over 500,000 childhood cancer survivors in the United States^1-3^ As a consequence of their cancer treatment, approximately 70% of survivors of childhood cancer develop clinically significant late effects that can result in premature mortality, increased morbidity, and reduced quality of life.^4-8^ Late effects often emerge during young adulthood and vary widely based on diagnosis and the types and doses of treatments received.^4^ They can include subsequent malignant neoplasms, chronic health conditions (e.g., cardiovascular and lung disease, renal dysfunction, musculoskeletal problems, endocrinopathies), psychosocial difficulties (e.g., posttraumatic stress, social impairment), and/or cognitive deficits, and require ongoing surveillance and management.^9^

Given the long-term late effects of treatment, it is critical that survivors of cancer demonstrate optimal self-management.^10^ Self-management is the interaction of health behaviors and related processes that patients and families engage in to care for a chronic condition.^11^ During adolescence and young adulthood self-management is increasingly important as they begin to form and consolidate critical health behaviors and assume increasing responsibility for their own health independent of parental influence.^12^ For adolescent and young adult (AYA) survivors of childhood cancer, self-management encompasses knowledge of cancer history and health risks, skills and confidence related to organization, planning, and/or administration of medical tasks, and engagement in cancer-related LTFU.^13-15^ Concerningly, AYA self-management is generally insufficient as evidenced by disease management skills, knowledge, health promoting behaviors, and engagement in follow-up care that is less than optimal and exacerbates health vulnerabilities of AYA survivors.^14, 16-23^

Poor engagement in follow-up care is one well-established vulnerability for AYA survivors. The Institute of Medicine (now the National Academy of Medicine) and Children’s Oncology Group (COG) recommend annual cancer-focused long-term follow-up (LTFU) visits across the life span.^5, 24, 25^ Such surveillance is needed to monitor for treatment-related late effects and adult malignancies that increase in incidence with age, and provide education for cancer prevention behaviors and psychosocial support, which may improve long-term survival.^13, 14, 26^ Specialized pediatric and adult cancer survivorship centers are critical for providing this care, especially given primary care providers are often unfamiliar with the risks and needs of childhood cancer survivors.^13-15^ Concerningly, LTFU attendance declines over time, particularly after the transition from pediatric to adult care, resulting in less than 30% of adult childhood cancer survivors receiving oncology LTFU.^16, 27-31^ In recent years, the NCI identified the combination of childhood cancer survivors facing higher risks for new cancers/morbidities and the decrease in LTFU care engagement as a health disparity and research priority.^17^

Given the importance of life-long LTFU oncology care, it is also essential that AYA become ready to transition to, and engage in, the adult health care system once they age out of pediatric care.^23^ Transition readiness is a gradual process, encompassing the purposeful, planned movement of AYA from pediatric-to adult-oriented health care.^32^ It consists of multifactorial indicators that the AYA and those in their system of support (family, providers) can begin, continue, and finish the transition process.^33, 34^ As outlined in our Social-Ecological Model of AYA Readiness to Transition (SMART),^35^ transition readiness is critical for AYA self-management in that it represents increasing maturity, knowledge, emotional readiness, and skills to sustain engagement in care. Increased transition readiness prior to transition is therefore expected to lead to higher engagement in pediatric-oriented care where transition planning will continue, indicating a potential transactional relationship between engagement in care and transition readiness.^33, 34^ However, advances in transition readiness research have been limited by historical reliance on cross-sectional data, lack of theory, and a paucity of valid measures of transition readiness, especially in pediatric oncology.^35, 36^ Additionally, few pediatric institutions offer standardized services for transferring childhood survivors to adult care,^37-41^ and only 8% of pediatric oncologists report using standardized transition readiness assessments to guide their care.^42^

To better understand trajectories of AYA survivor self-management and transition readiness, we created the AYA Tracking Engagement and Management Skills (AYA TEAMS) longitudinal observational cohort consisting of AYA survivors of childhood cancer enrolled prior to the transition to adult care. Specifically, the AYA TEAMS study aims to: 1) validate a novel and comprehensive measure of transition readiness, the Transition Readiness Inventory (TRI); 2) test transactional relationships among transition readiness, self-management skills, and engagement in pediatric LTFU; and 3) identify predictors of engagement in adult LTFU care. Here we describe the cohort profile of the AYA TEAMS study, including design and methods, participant recruitment, and the demographic and clinical characteristics of the baseline sample. This cohort will serve as the basis from which we will test the aims of our longitudinal study in subsequent analysis and papers.

### COHORT DESCRIPTION

#### Study Design

The AYA TEAMS study enrolled AYA survivors of childhood cancer from 3 large pediatric cancer centers across the United States: The Children’s Hospital of Philadelphia (CHOP, Northwest), Cincinnati Children’s Hospital Medical Center (CCHMC, Midwest), and Children’s Hospital Los Angeles (CHLA, West). Participants who were not transferred to adult care during the study completed three annual assessments (T1, T2, T3). If they were transferred between T1 and T3, they followed a modified schedule of assessments that included: 1) assessment after their last pediatric visit (designated transfer assessment #1, or Tr1) and 2) a 15-month follow-up (Tr15) to assess initial engagement in adult-focused LTFU after transfer. Caregivers of enrolled AYA were also asked to complete a single survey at baseline. A subset of AYA and caregivers from CHOP completed the TRI 1 week (5-15 days) after baseline (T1) to evaluate test-retest reliability. See Figure 1 for study data collection time points. AYA completed online surveys at each timepoint, and medical history and attendance at LTFU care were abstracted from each patient’s electronic health record (EHR). All data were collected and managed using REDCap electronic data capture tools hosted at CHOP.^43^ The CHOP IRB served as the central IRB and reviewed and approved this study protocol (IRB 17-013782).

**Fig 1.**
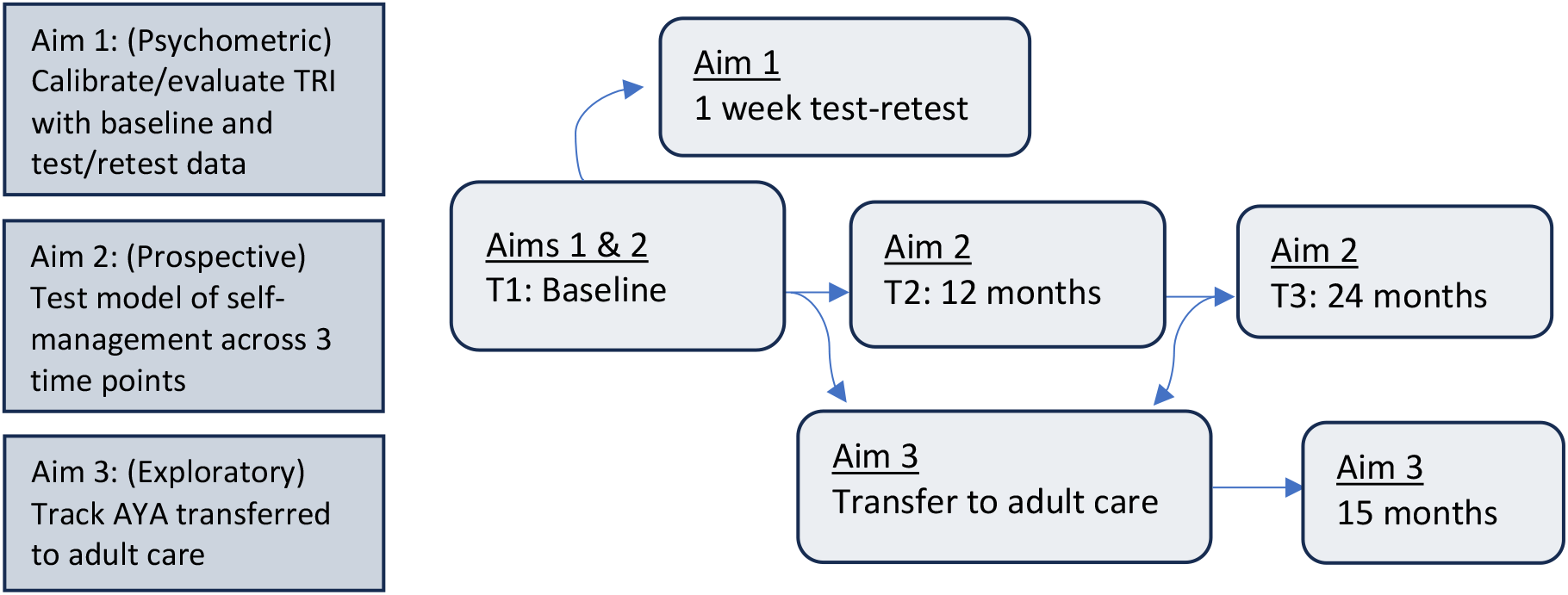
Study Aims and Points of Data Collection AYA= Adolescents and Young Adults; TRI=Transition Readiness Inventory.

#### Participant eligibility and recruitment

Inclusion criteria for AYA included: 1) 16 to 25 years-old and a survivor of childhood cancer; 2) at least five years post diagnosis; 3) at least two years from end of treatment; 4) cognitively capable of completing study procedures; 5) English-speaking; 6) engaged within the enrolling pediatric hospital (any department/specialty) within the last 18 months; and 7) living in the U.S. Exclusion criteria included 1) already transferred to adult oncology care, 2) deceased or 3) provider did not provide approval to approach. Eligibility for caregivers included being a parent or primary caregiver of a non-married AYA participant and English or Spanish speaking. We attempted to enroll one parent/caregiver for each AYA enrolled, but AYA could elect to decline caregiver participation/contact. Eligible AYA were identified at the site level via EHR review and survivorship clinic lists and verified with oncology clinicians. AYA and caregivers were approached in clinic or via telephone. Caregivers and AYA over 18 years provided consent (in person or over the phone). AYA under 18 years provided assent and a parent or legal guardian provided consent, with AYA providing consent after they turned 18. AYA were passively withdrawn from the cohort and considered lost to follow-up if they completed less than half of the baseline (T1) survey.

#### Study Measures

Clinical characteristics including cancer diagnosis (type of cancer, age at diagnosis, relapse, second cancer), treatment history (including modalities and time off treatment), and late effects were collected via EHR review. Late effects, including current cancer-related health problems and future risks related to treatment, were recorded based on documentation in the most recent oncology clinic note. Eighteen categories of late effects were abstracted: endocrine (including growth, thyroid and diabetes), cognitive, dermatological, vision, hearing, dental, cardiac, respiratory, kidney, gastrointestinal, reproductive, bladder/urinary, immunological, bone/joint, psychological, second cancers, neurological (including headaches, seizures and balance problems), and liver problems. If it was unclear whether a health problem was associated with past cancer and/or treatment history, it was reviewed with an oncologist to verify. Intensity of Treatment Ratings (ITR) were rated using standardized procedures^44^ from 1 (least intense) to 4 (most intense). All ratings were verified by an oncology clinician (survivorship physician or advanced practice provider), with discrepancies discussed and resolved.

Demographic characteristics, including age and sex at birth were recorded from the EHR. Using participant addresses, neighborhood median income and low-income data were extracted from the U.S. Bureau of the Census 2020 Census Summary at the Census Tract (CT) level. A comprehensive battery of self-reported measures assessed additional demographic characteristics, transition readiness components, potential predictors of transition readiness, self-management skills, and engagement in LTFU (see Table 1). After the onset of COVID-19, additional measures were added to assess COVID-19 exposures and impact (May 2020), risk and resiliency (June 2020) and COVID-19 vaccine hesitancy (Nov 2020) to use as potential covariates for planned analysis. REDCap survey links were distributed to participants via email and text message. See Figure 2 for the proposed transactional model among constructs.

**Table 1:**
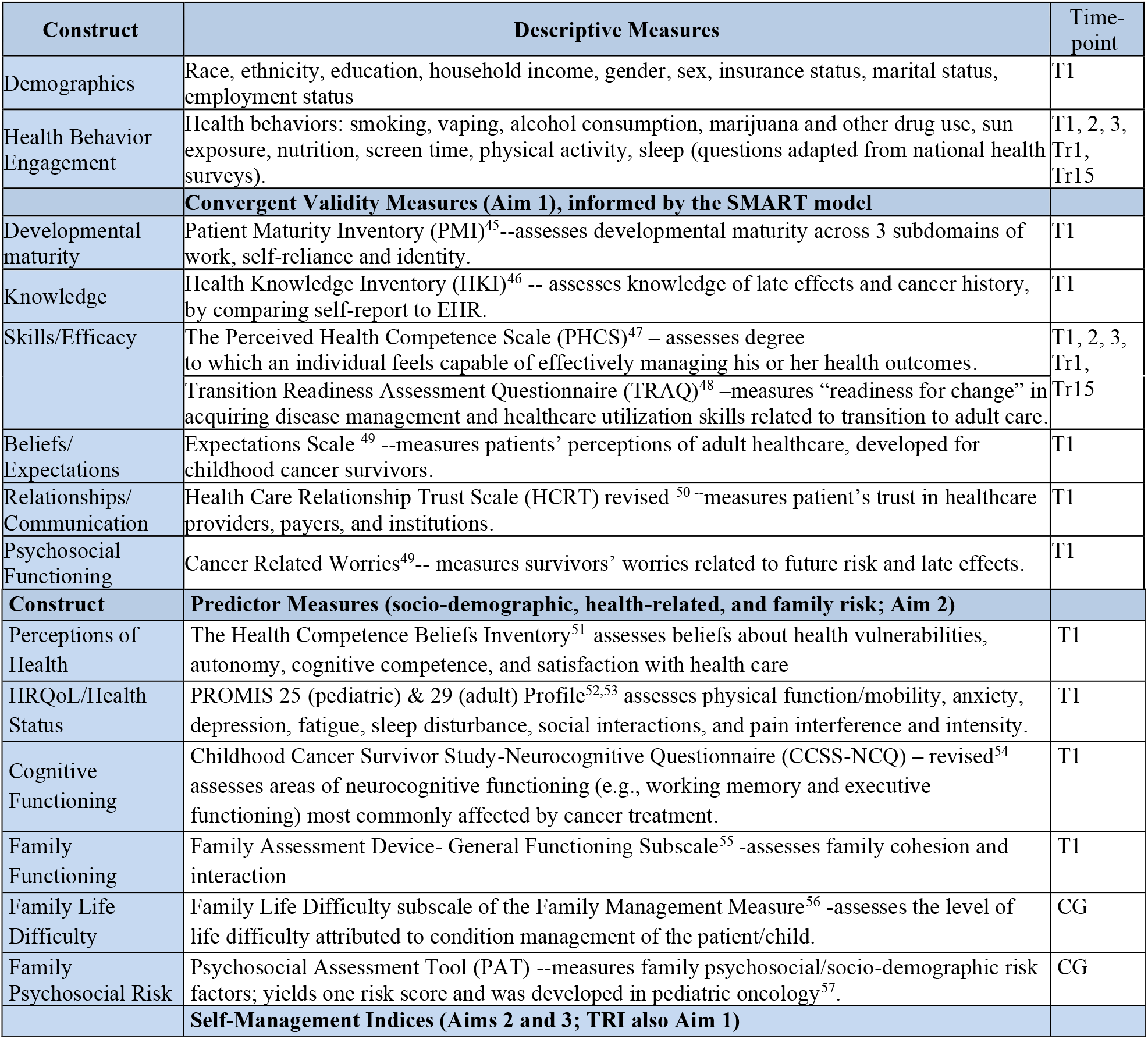
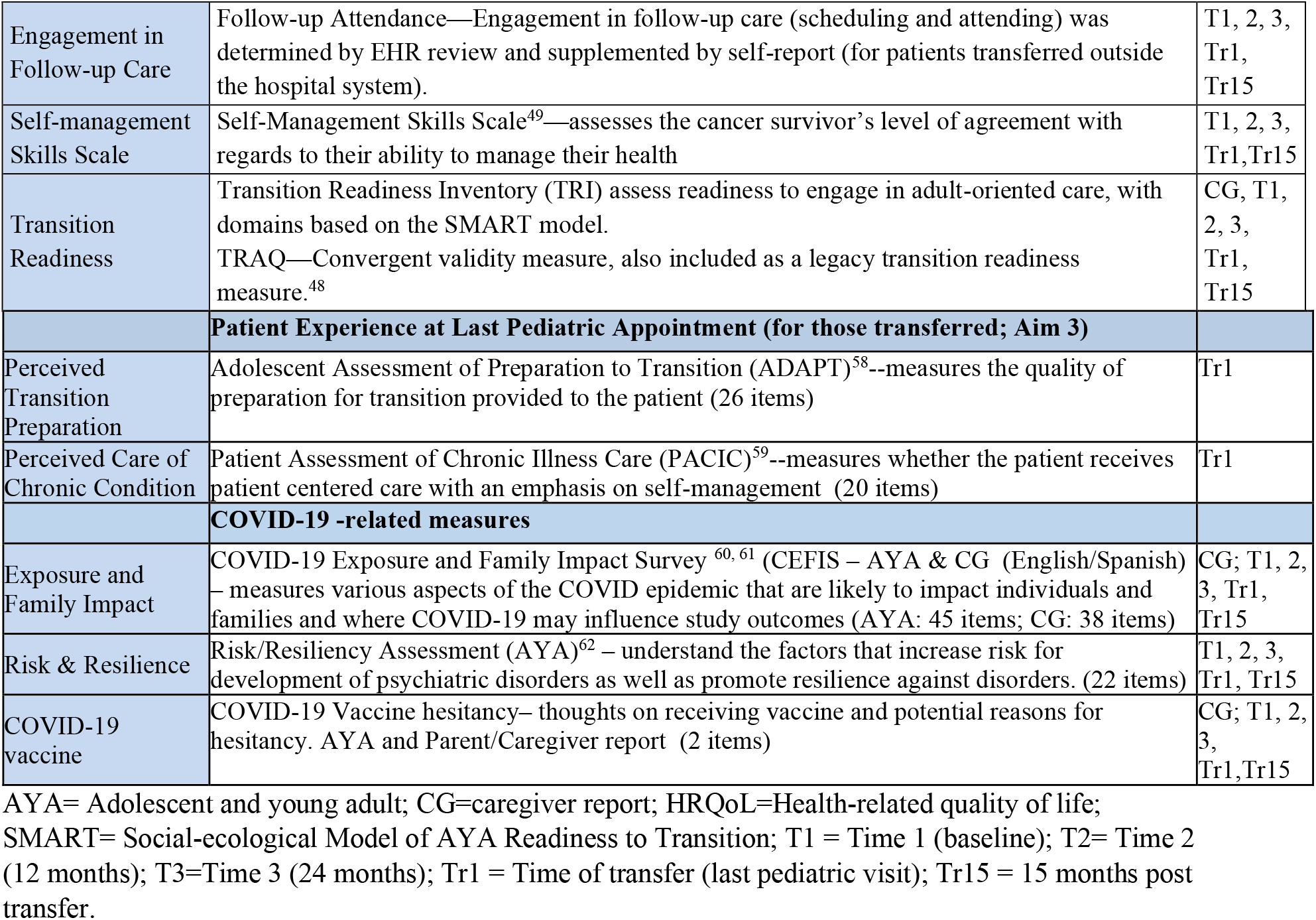
Self-reported Survey Measures.

| Construct | Descriptive Measures | Time-point |
| --- | --- | --- |
| Demographics | Race, ethnicity, education, household income, gender, sex, insurance status, marital status, employment status | T1 |
| Health Behavior Engagement | Health behaviors: smoking, vaping, alcohol consumption, marijuana and other drug use, sun exposure, nutrition, screen time, physical activity, sleep (questions adapted from national health surveys). | T1, 2, 3, Tr1, Tr15 |
|  | **Convergent Validity Measures (Aim 1), informed by the SMART model** |  |
| Developmental maturity | Patient Maturity Inventory (PMI)<sup>45</sup>--assesses developmental maturity across 3 subdomains of work, self-reliance and identity. | T1 |
| Knowledge | Health Knowledge Inventory (HKI)<sup>46</sup> -- assesses knowledge of late effects and cancer history, by comparing self-report to EHR. | T1 |
| Skills/Efficacy | The Perceived Health Competence Scale (PHCS)<sup>47</sup> – assesses degree to which an individual feels capable of effectively managing his or her health outcomes.<br>Transition Readiness Assessment Questionnaire (TRAQ)<sup>48</sup> –measures “readiness for change” in acquiring disease management and healthcare utilization skills related to transition to adult care. | T1, 2, 3, Tr1, Tr15 |
| Beliefs/Expectations | Expectations Scale <sup>49</sup> --measures patients’ perceptions of adult healthcare, developed for childhood cancer survivors. | T1 |
| Relationships/Communication | Health Care Relationship Trust Scale (HCRT) revised <sup>50</sup>--measures patient’s trust in healthcare providers, payers, and institutions. | T1 |
| Psychosocial Functioning | Cancer Related Worries<sup>49</sup>-- measures survivors’ worries related to future risk and late effects. | T1 |
| **Construct** | **Predictor Measures (socio-demographic, health-related, and family risk; Aim 2)** |  |
| Perceptions of Health | The Health Competence Beliefs Inventory<sup>51</sup> assesses beliefs about health vulnerabilities, autonomy, cognitive competence, and satisfaction with health care | T1 |
| HRQoL/Health Status | PROMIS 25 (pediatric) & 29 (adult) Profile<sup>52,53</sup> assesses physical function/mobility, anxiety, depression, fatigue, sleep disturbance, social interactions, and pain interference and intensity. | T1 |
| Cognitive Functioning | Childhood Cancer Survivor Study-Neurocognitive Questionnaire (CCSS-NCQ) – revised<sup>54</sup> assesses areas of neurocognitive functioning (e.g., working memory and executive functioning) most commonly affected by cancer treatment. | T1 |
| Family Functioning | Family Assessment Device- General Functioning Subscale<sup>55</sup> -assesses family cohesion and interaction | T1 |
| Family Life Difficulty | Family Life Difficulty subscale of the Family Management Measure<sup>56</sup> -assesses the level of life difficulty attributed to condition management of the patient/child. | CG |
| Family Psychosocial Risk | Psychosocial Assessment Tool (PAT) --measures family psychosocial/socio-demographic risk factors; yields one risk score and was developed in pediatric oncology<sup>57</sup>. | CG |
|  | **Self-Management Indices (Aims 2 and 3; TRI also Aim 1)** |  |
| Engagement in Follow-up Care | Follow-up Attendance—Engagement in follow-up care (scheduling and attending) was determined by EHR review and supplemented by self-report (for patients transferred outside the hospital system). | T1, 2, 3, Tr1, Tr15 |
| Self-management Skills Scale | Self-Management Skills Scale<sup>49</sup>—assesses the cancer survivor’s level of agreement with regards to their ability to manage their health | T1, 2, 3, Tr1, Tr15 |
| Transition Readiness | Transition Readiness Inventory (TRI) assess readiness to engage in adult-oriented care, with domains based on the SMART model.<br>TRAQ—Convergent validity measure, also included as a legacy transition readiness measure.<sup>48</sup> | CG, T1, 2, 3, Tr1, Tr15 |
| **Patient Experience at Last Pediatric Appointment (for those transferred; Aim 3)** |  |  |
| Perceived Transition Preparation | Adolescent Assessment of Preparation to Transition (ADAPT)<sup>58</sup>--measures the quality of preparation for transition provided to the patient (26 items) | Tr1 |
| Perceived Care of Chronic Condition | Patient Assessment of Chronic Illness Care (PACIC)<sup>59</sup>--measures whether the patient receives patient centered care with an emphasis on self-management (20 items) | Tr1 |
| **COVID-19 -related measures** |  |  |
| Exposure and Family Impact | COVID-19 Exposure and Family Impact Survey<sup>60, 61</sup> (CEFIS – AYA & CG (English/Spanish) – measures various aspects of the COVID epidemic that are likely to impact individuals and families and where COVID-19 may influence study outcomes (AYA: 45 items; CG: 38 items) | CG; T1, 2, 3, Tr1, Tr15 |
| Risk & Resilience | Risk/Resiliency Assessment (AYA)<sup>62</sup> – understand the factors that increase risk for development of psychiatric disorders as well as promote resilience against disorders. (22 items) | T1, 2, 3, Tr1, Tr15 |
| COVID-19 vaccine | COVID-19 Vaccine hesitancy– thoughts on receiving vaccine and potential reasons for hesitancy. AYA and Parent/Caregiver report (2 items) | CG; T1, 2, 3, Tr1, Tr15 |

AYA= Adolescent and young adult; CG=caregiver report; HRQoL=Health-related quality of life; SMART= Social-ecological Model of AYA Readiness to Transition; T1 = Time 1 (baseline); T2= Time 2 (12 months); T3=Time 3 (24 months); Tr1 = Time of transfer (last pediatric visit); Tr15 = 15 months post transfer.

**Fig 2.**
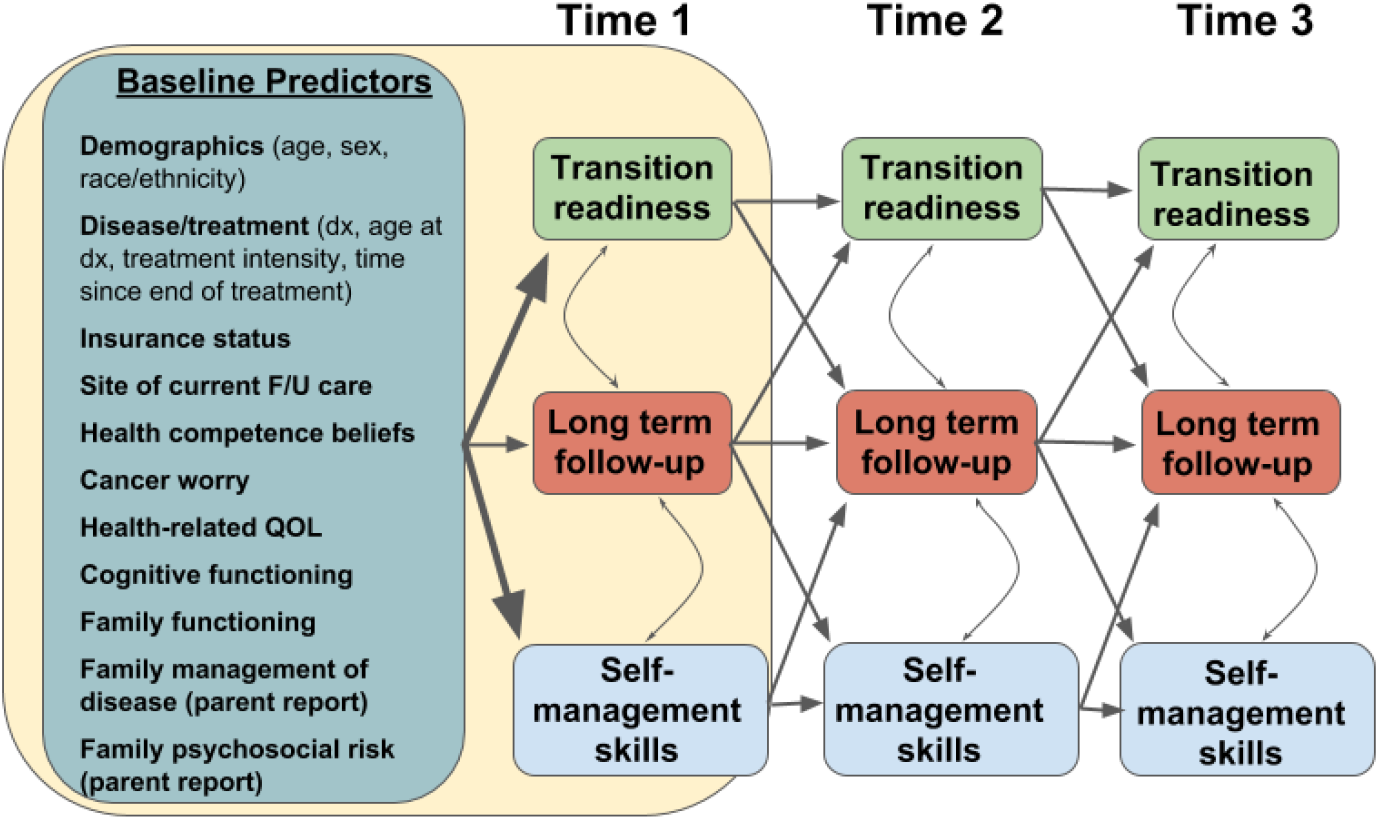
Proposed transactional model Note: Dx=diagnosis; F/U = follow-up; QOL=quality of life

#### Statistical Analysis

Descriptive statistics (means, SD, ranges, frequencies) for baseline sociodemographic and clinical characteristics were calculated to describe the AYA TEAMS baseline cohort. These results were examined for the sample in aggregate and by each study site. Differences in clinical and demographic characteristics between study sites were examined via Chi square and Analysis of Variance, with Bonferroni post-hoc adjustments. We also conducted an exploratory analysis to examine differences between enrolled AYA included in the cohort and those who never completed baseline based on age, sex, race/ethnicity, cancer diagnosis type, caregiver enrollment and site using Chi square and t-tests. All analyses were conducted in SPSS version 28.0.

#### Patient and public involvement

Patients were not involved in the design of this research or the writing or editing of this document. Study updates and results were disseminated to participants via emailed newsletters.

#### Participant Recruitment

In total, 785 AYA were approached, 709 (90.3%) were enrolled, and of those 587 (82.8%) participants completed at least half of the baseline measures survey between December 2019 and October 2022 and were included in the cohort (See Fig 3). For caregivers, of the 709 AYA enrolled, 518 (73.1%) caregivers were approached and of those 480 (92.7%) enrolled (See Fig 4). Across sites, the majority of AYA were recruited in person at clinic appointments (n=413, 58.3%), with the remainder recruited remotely over the phone. For CHLA and CCHMC, phone-based recruitment was more prevalent (66% and 73%, respectively) largely due to COVID-19 pandemic restrictions (starting March 2020) and clinic preferences for limiting in-person patient contact. For phone-based recruitment we attempted to contact patients around the time of their oncology clinic visits.

**Fig 3:**
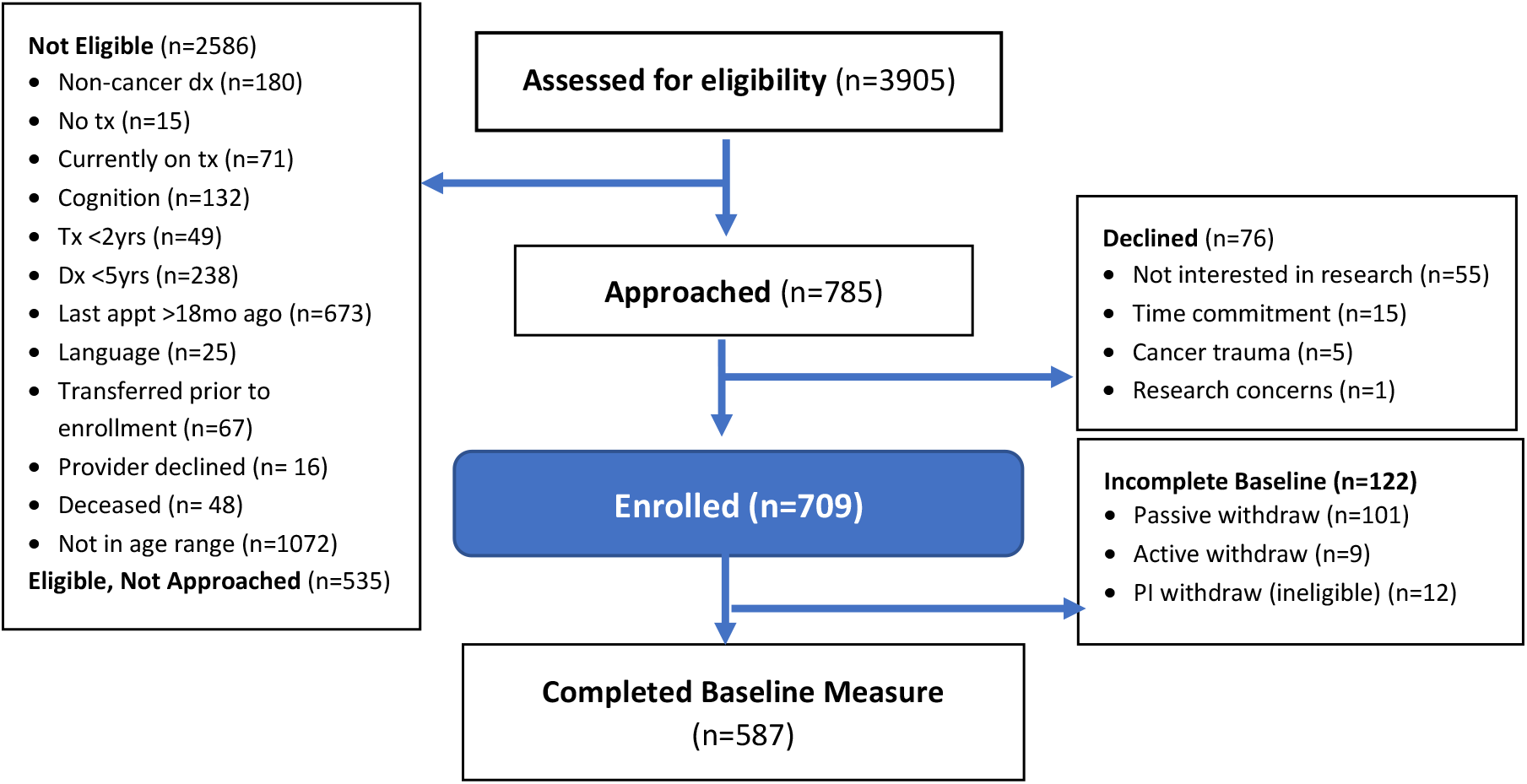
AYA Participant Flow Diagram

**Fig 4:**
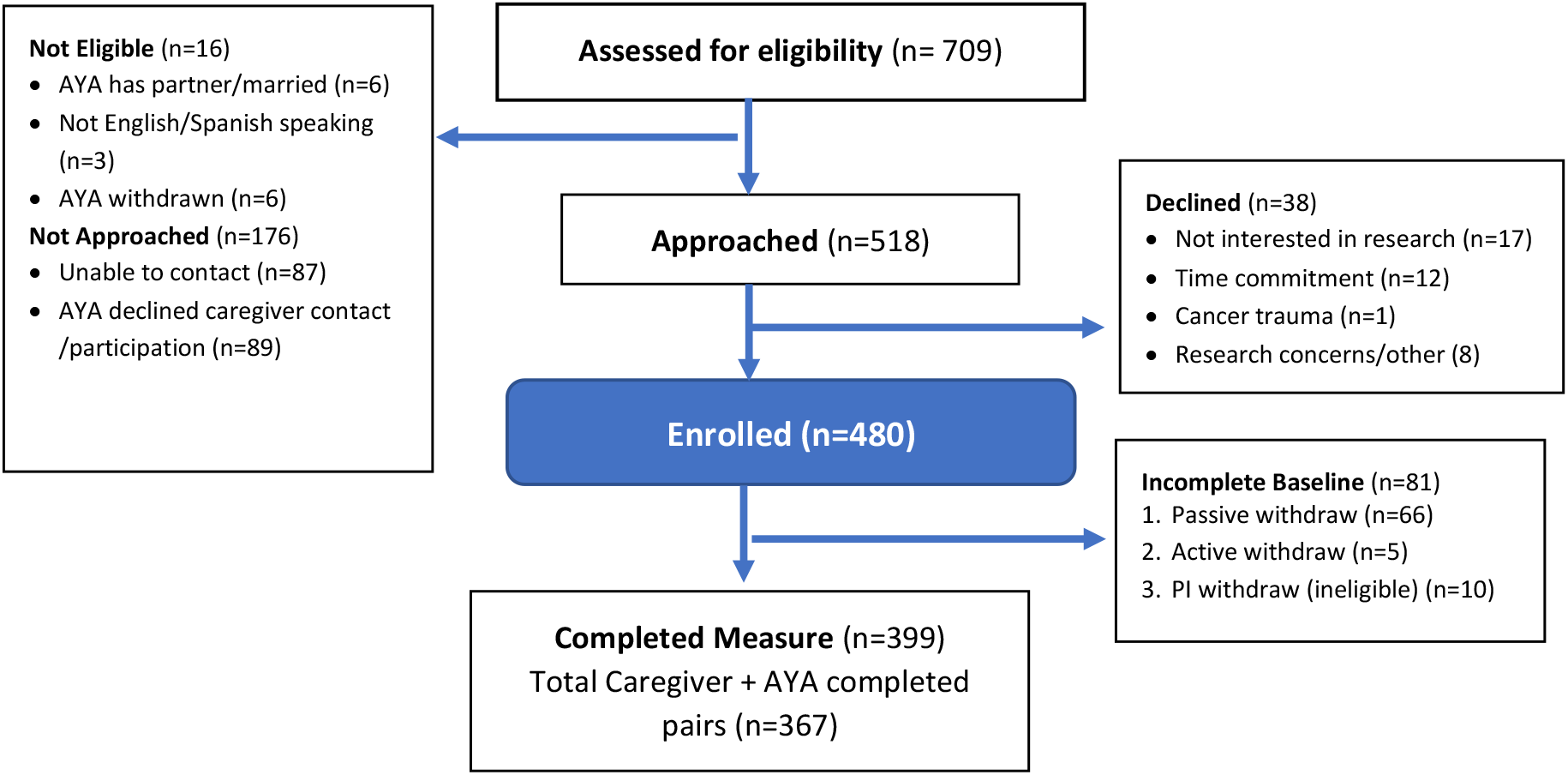
Caregiver Participant Flow Diagram

#### Participant characteristics

##### AYA

Clinical and demographic characteristics of our baseline cohort at each site and combined can be found in Tables 2 and 3. The cohort was on average 7.3 years old at the time of diagnosis and 10.5 years off treatment and about half the sample (53%) were survivors of leukemia or lymphoma. Almost all AYA had been previously seen in a dedicated cancer survivorship clinic (95%), with more than half (n=336, 57.2%) enrolled on the same day of their oncology clinic visit, and 75.8% of the sample enrolled within 1 month. More than one-third of AYA reported being from a racial or ethnic minoritized group (38%) and 52% identified as female.

**Table 2:**
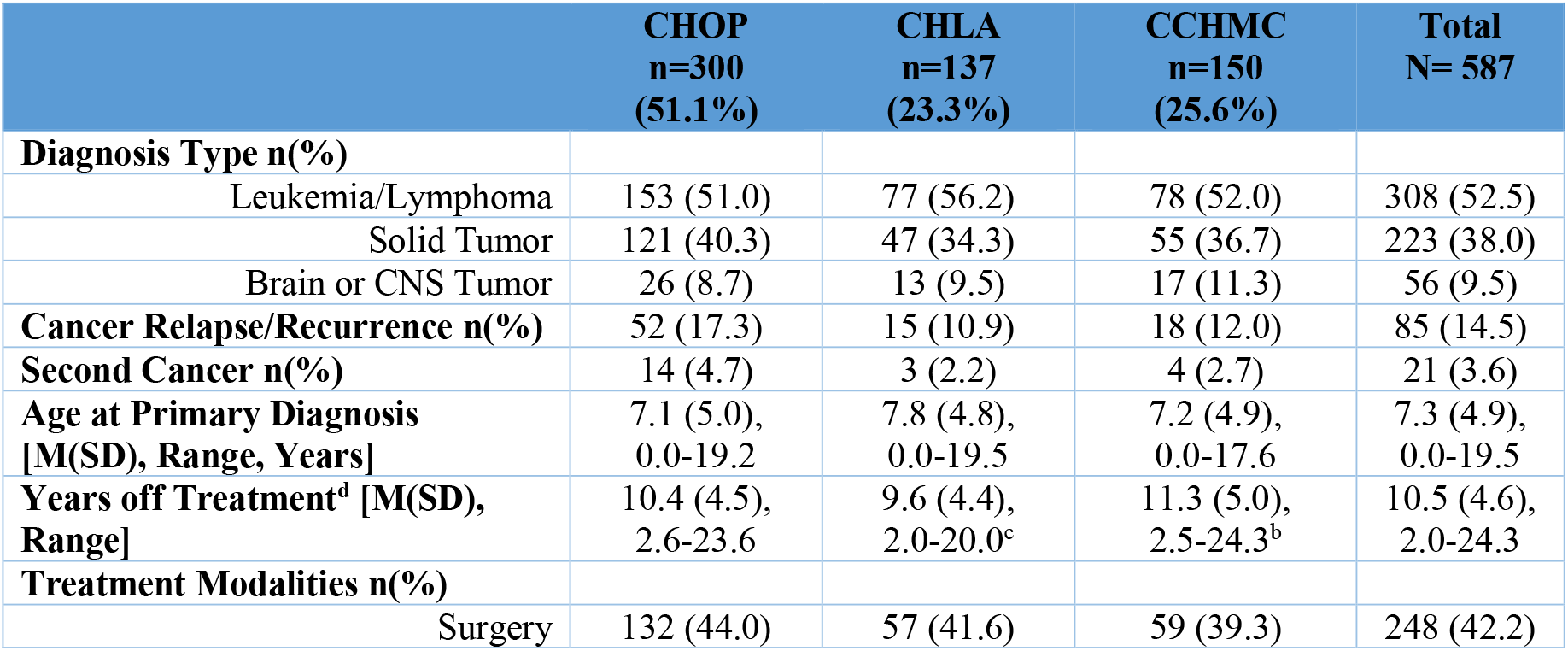
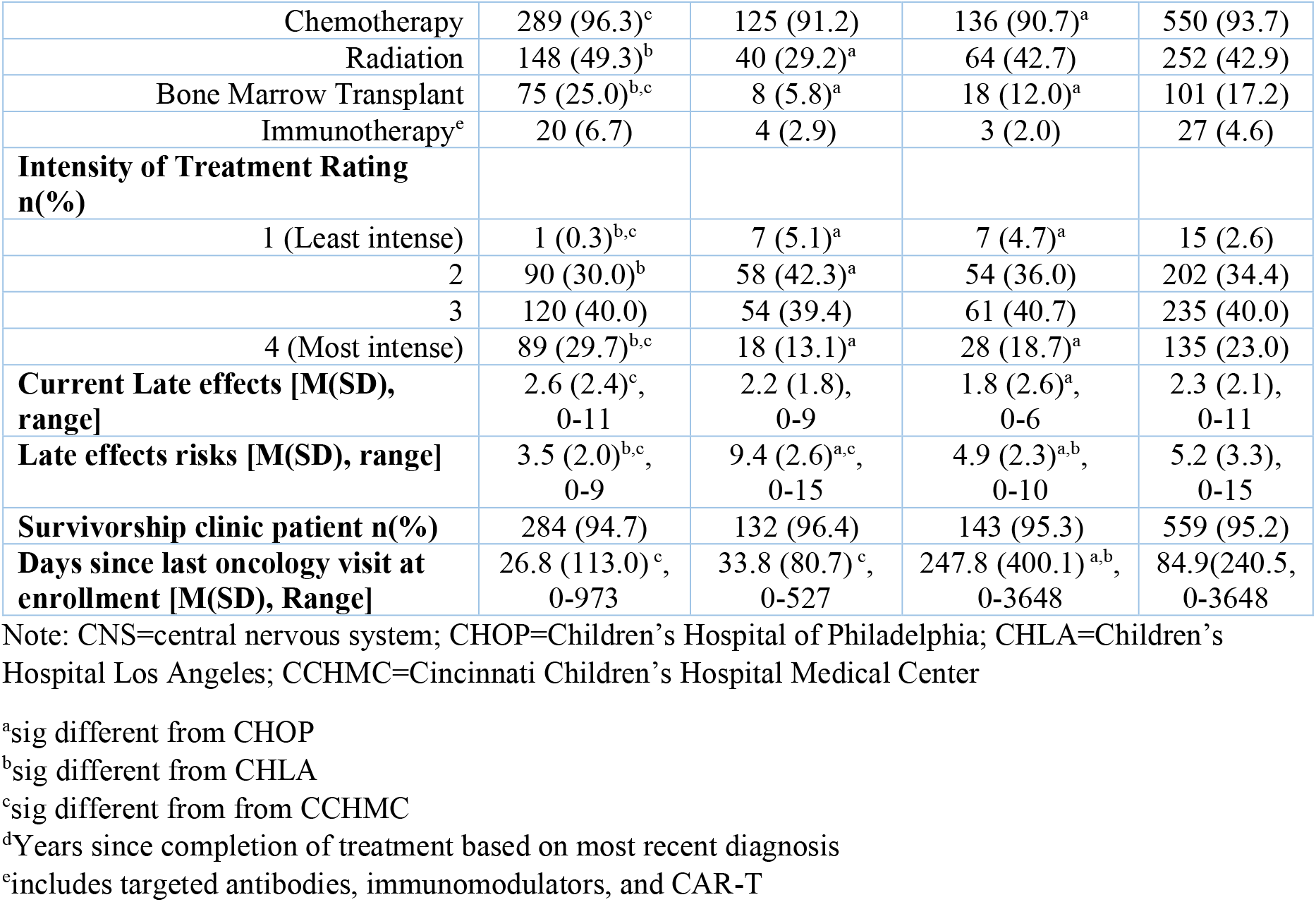
AYA Clinical Characteristics.

**Table 3:**
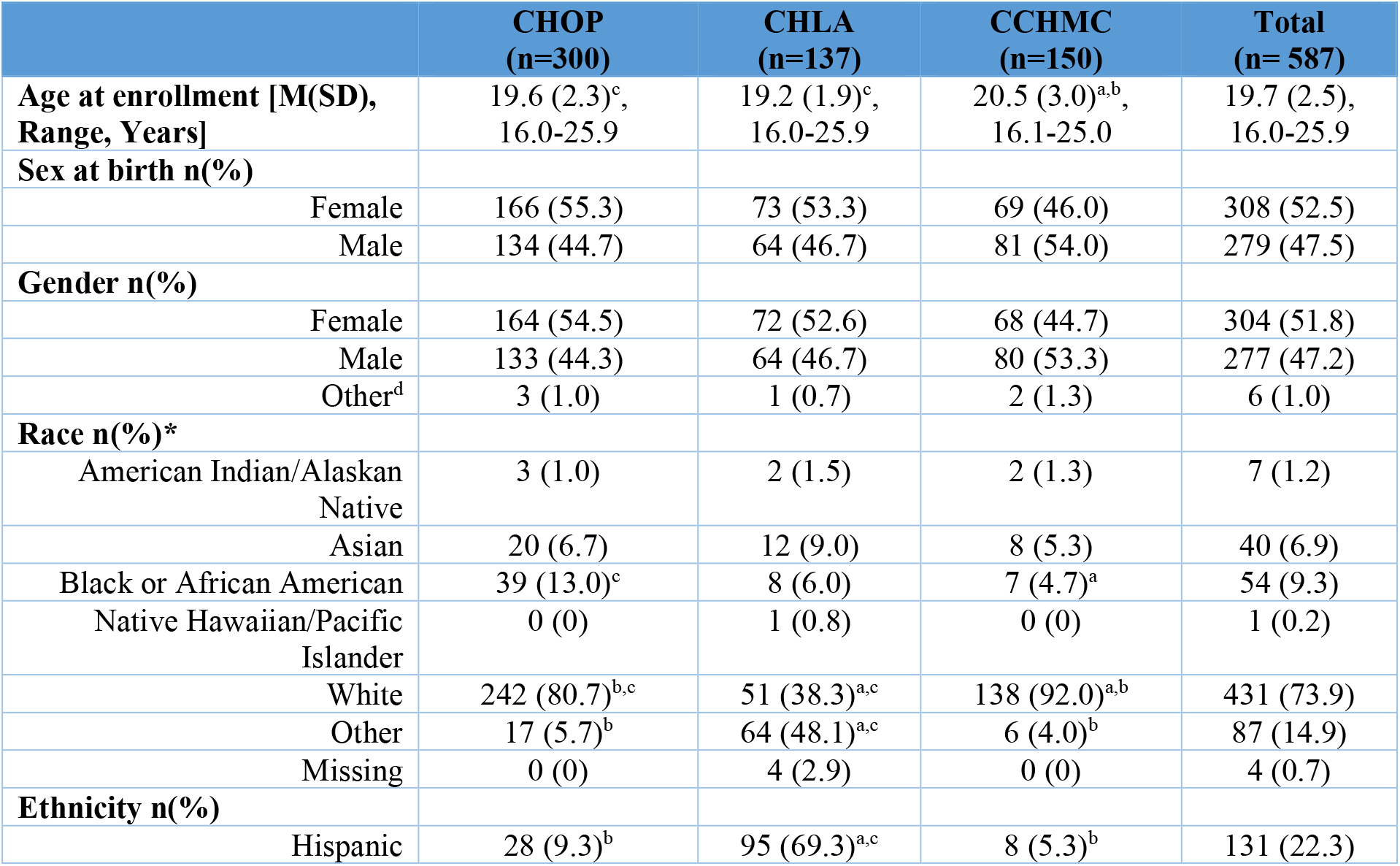
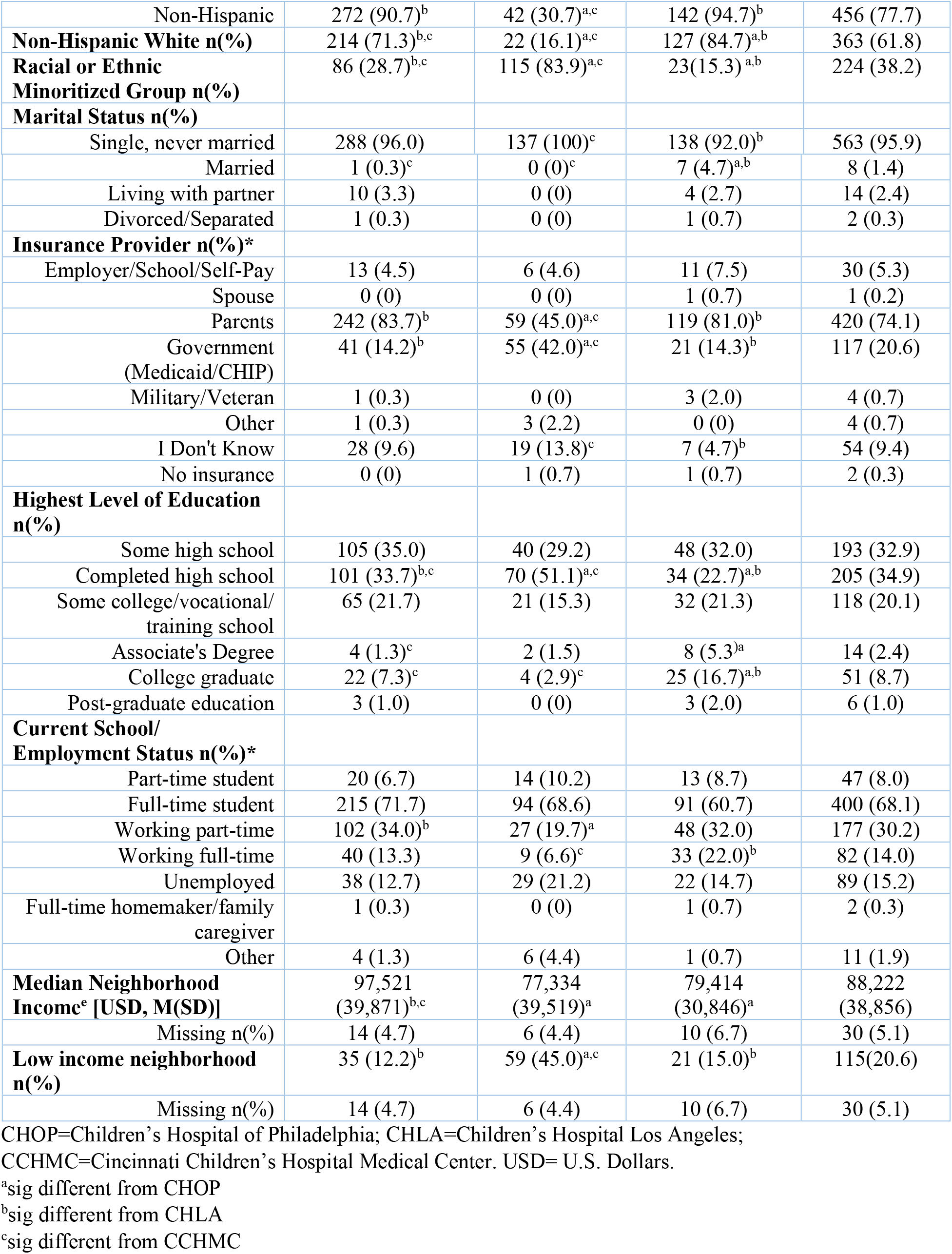
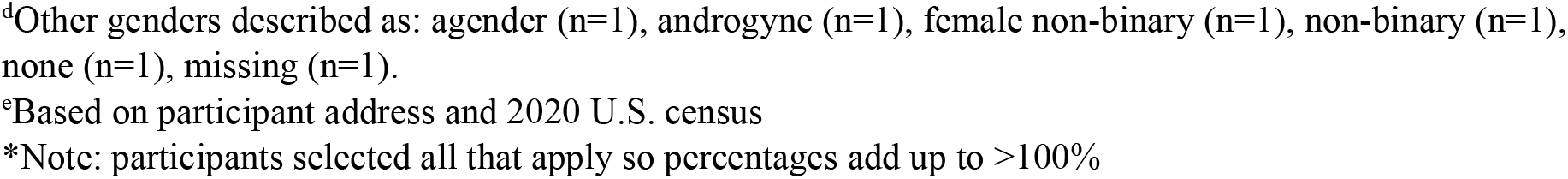
AYA Demographic Characteristics.

##### Caregivers

In total, 399 caregivers completed the caregiver survey. Most caregivers were mothers (90%) and completed the English survey (94%). See Table 4 for detailed caregiver demographics.

**Table 4:** Caregiver Demographic Characteristics.

|  | CHOP<br>(n=259) | CHLA<br>(n=69) | CCHMC<br>(n=71) | Total<br>(n= 399) |
| --- | --- | --- | --- | --- |
| **Age at survey [(M/SD), range]** | 50.9 (6.0),<br>36-64<sup>b</sup> | 48.3 (7.3),<br>34-75<sup>a</sup> | 49.7 (5.4),<br>38-62 | 50.3 (6.3),<br>34-75 |
| Missing n(%) | 2 (0.8) | 0 (0) | 0 (0) | 2 (0.5) |
| **Gender n(%)** |  |  |  |  |
| Female | 235 (90.7) | 66 (95.7) | 65 (91.5) | 366 (91.7) |
| Male | 24 (9.3) | 3 (4.3) | 6 (8.5) | 33 (8.3) |
| **Relationship to AYA n(%)** |  |  |  |  |
| Mother | 232 (89.6) | 63 (91.3) | 64 (90.1) | 359 (90.0) |
| Father | 24 (9.3) | 3 (4.3) | 6 (8.5) | 33 (8.3) |
| Grandparent | 3 (1.2) | 2 (2.9) | 1 (1.4) | 6 (1.5) |
| Other | 0 (0) | 1 (1.4) | 0 (0) | 1 (0.3) |
| **Race n(%)\*** |  |  |  |  |
| American Indian/Alaskan Native | 3 (1.2) | 1 (1.5) | 0 (0) | 4 (1.0) |
| Asian | 14 (5.4) | 4 (6.1) | 3 (4.2) | 21 (5.3) |
| Black or African American | 21 (8.1) | 4 (6.1) | 1 (1.4) | 26 (6.6) |
| Native Hawaiian/Pacific Islander | 1 (0.4) | 0 (0) | 0 (0) | 1 (0.3) |
| White | 219 (84.6)<sup>b,c</sup> | 33 (50.0)<sup>a,c</sup> | 69 (97.2)<sup>a,b</sup> | 321 (81.1) |
| Other | 10 (3.9)<sup>b</sup> | 24 (34.8)<sup>a,c</sup> | 0 (0)<sup>b</sup> | 34 (8.6) |
| Missing | 0 (0) | 3 (4.3) | 0 (0) | 3 (0.8) |
| **Ethnicity n(%)** |  |  |  |  |
| Hispanic | 14 (5.4)<sup>b</sup> | 46 (66.7)<sup>a,c</sup> | 2 (2.8)<sup>b</sup> | 62 (15.5) |
| Non-Hispanic | 245 (94.6)<sup>b</sup> | 23 (33.3)<sup>a,c</sup> | 69 (97.2)<sup>b</sup> | 337 (84.5) |
| **Non-Hispanic White n(%)** | 207 (79.9)<sup>b,c</sup> | 15 (21.7)<sup>a,c</sup> | 65 (91.5)<sup>a,b</sup> | 287 (71.9) |
| **Racial or Ethnic Minoritized Group n(%)** | 52 (20.1)<sup>b,c</sup> | 54 (78.3)<sup>a,c</sup> | 6 (8.5)<sup>a,b</sup> | 112 (28.7) |
| **Survey language n(%)** |  |  |  |  |
| English | 256 (98.8)<sup>b</sup> | 49 (71.0)<sup>a,c</sup> | 71 (100)<sup>b</sup> | 376 (94.2) |
| Spanish | 3 (1.2)<sup>b</sup> | 20 (29.0)<sup>a,c</sup> | 0 (0)<sup>b</sup> | 23 (5.8) |
| **Annual Household income [USD, n(%)]<sup>d</sup>** |  |  |  |  |
| Less than 25,000 | 7 (2.7)<sup>b</sup> | 17 (24.6)<sup>a,c</sup> | 4 (5.6)<sup>b</sup> | 28 (7.1) |
| 25,000-49,000 | 25 (9.7)<sup>b</sup> | 22 (31.9)<sup>a,c</sup> | 6 (8.5)<sup>b</sup> | 53 (13.3) |
| 50,000-99,000 | 72 (27.8)<sup>b</sup> | 6 (8.7)<sup>a,c</sup> | 25 (35.2)<sup>b</sup> | 103 (25.8) |
| 100,000 or more | 146 (56.4)<sup>b</sup> | 21 (30.4)<sup>a</sup> | 35 (49.3) | 202 (50.6) |
| Don't know | 7 (2.7) | 3 (4.3) | 1 (1.4) | 11 (2.8) |
| Missing | 2 (0.8) | 0 (0) | 0 (0) | 2 (0.5) |
| **Marital Status n(%)** |  |  |  |  |
| Single | 17 (6.6)<sup>b</sup> | 12 (17.4)<sup>a</sup> | 4 (5.6) | 33 (8.3) |
| Married/Partnered | 198 (76.7) | 48 (69.6) | 58 (81.7) | 304 (76.4) |
| Separated/Divorced | 39 (15.1) | 5 (7.2) | 6 (8.5) | 50 (12.6) |
| Widowed | 4 (1.6) | 4 (5.8) | 2 (2.8) | 10 (2.5) |
| Other | 0 (0) | 0 (0) | 1 (1.4) | 1 (0.3) |
| Missing | 1 (0.4) | 0 (0) | 0 (0) | 1 (0.3) |
| **Highest Education n(%)** |  |  |  |  |
| Less than high school | 2 (0.8)<sup>b</sup> | 15 (21.7)<sup>a,c</sup> | 2 (2.9)<sup>b</sup> | 19 (4.8) |
| Finished high school/GED | 33 (12.8) | 16 (23.2) | 12 (17.1) | 61 (15.4) |
| Started college/trade school | 13 (12.0) | 13 (18.8) | 9 (12.9) | 53 (13.4) |
| Finished college/trade school | 125 (48.4) | 22 (31.9) | 32 (45.7) | 179 (45.1) |
| Started master's or doctorate program | 13 (5.0) | 1 (1.4) | 2 (2.9) | 16 (4.0) |
| Finished master's or doctorate program | 60 (23.3) | 7 (10.1) | 18 (25.7) | 85 (21.4) |
| Missing | 1 (0.4) | 0 (0) | 1 (1.4) | 2 (0.5) |

CHOP=Children's Hospital of Philadelphia; CHLA=Children's Hospital Los Angeles; CCHMC=Cincinnati Children's Hospital Medical Center. USD= U.S. Dollars.
<sup>a</sup>sig different from CHOP
<sup>b</sup>sig different from CHLA
<sup>c</sup>sig different from CCHMC
<sup>d</sup>Based on caregiver self-report
\*Note: participants selected all that apply so percentages add up to >100%

### FINDINGS TO DATE

#### Baseline attrition

Eligible AYA who consented, but did not complete at least half of the baseline survey and excluded from the cohort (n=110, 15.5%) were more likely male (69.1% vs. 47.5%, X^2^ =17.23, p<.001), non-white and/or Hispanic as indicated in the EHR (63.9% vs 49.1%, X^2^=7.1, p=.008), and more likely to enroll alone without a caregiver (71.0% vs. 54.9%, X^2^=9.31, p=.002) compared to AYA who were included in the cohort (n=587). CHLA had the highest rates of dropout prior to completing baseline (25.9% compared to CHOP 12.0% and CCHMC 12.3%, X^2^=10.6, p<.001). No differences in baseline attrition were identified based on age at enrollment (19.8 vs. 19.7 years, t=0.35, p=.76) or diagnosis type (X^2^=1.14, p=.57).

#### Cohort clinical characteristics across sites

Among AYA included in the cohort (n=587), no site differences across cancer types (X^2^=2.2, p=.69), cancer relapse (X^2^=4.1, p=.13), second cancer occurrence (X^2^=2.2, p=.34), surgery (X^2^=0.9, p=.63), or age at diagnosis (F=1.01, p=.37) were found. Site differences in other modalities of treatment emerged, with CHOP having a higher percentage of patients receiving chemotherapy (compared to CCHMC X^2^=7.3, p=.03), radiation (compared to CHLA, X^2^=15.6, p<.001), and bone marrow transplant (compared to both sites, X^2^=28.1, p<.001). Overall, AYA recruited from CHOP had a highest intensity of treatment, with the fewest participants receiving a score of 1 (minimally intensive) and the highest percentage of participants with an ITR of 4 (most intensive, X^2^=29.1, p<.001). AYA from CHOP had a greater number of current late effects compared to AYA from CCHMC (F=7.3, p<.001), whereas CHLA had the greatest number of late effects risks documented in the EHR (F=328.0, p<.001). AYA from CCHMC were off treatment for longer compared to those recruited from CHLA (F=4.7, p=.010), and were enrolled further out from their last clinic visit compared to CHOP and CHLA participants (F=54.7, p<.001).

#### Cohort demographics across sites

Significant differences in demographic variables were observed across sites. AYA at CCHMC were slightly older (F=12.1, p<.001), had achieved a higher level of education (X^2^=48.6, p<.001), and were more likely to be married (X^2^=22.1, p=.001) compared to AYA from CHOP and CHLA. CHLA and CCHMC participants were from neighborhoods with lower average median income than CHOP participants (F=18.0, p<.001), with CHLA participants more likely living in low-income neighborhoods than AYA from both other sites (X^2^=62.6, p<.001). Racial and ethnic diversity differed across all sites, with CHLA having the greatest percentage from racial or ethnically minoritized groups (REMG, X^2^=166.3, p<.001). AYA at CHLA were also less likely to be covered under their parent’s health insurance (X^2^=75.2, p<.001) and more likely to have public insurance (X^2^=47.4, p<.001). There were no differences in AYA sex distribution across sites (X^2^=3.5, p=.17). Demographic differences in the caregiver sample reflected that of AYA, with caregivers from CHLA more likely to identify as from REMG (X^2^=107.8, p<.001) and reporting lower household income (X^2^=77.9, p<.001) compared to the other two sites. Caregivers at CHLA were younger (F=5.4, p=.005), more likely to be single (X2 = 22.1, p=.015), with a trend for being less likely to have graduated college (X^2^=5.9, p=.052) compared to CHOP caregivers. Caregivers from CHLA were less likely to have finished high school compared to both CHOP and CCHMC caregivers (X^2^=53.5, p<.001).

#### Discussion

The AYA TEAMS cohort is a medically and demographically diverse cohort of AYA survivors of childhood cancer, including AYA across all cancer types and treatment modalities. According to recent (2022) U.S. childhood cancer statistics for survivors under age 30, approximately 48% are female, 51% are survivors of leukemia or lymphoma, 31% are survivors of a solid tumor and 18% are survivors of CNS tumor.^63^ The lower proportion of CNS tumor survivors in our cohort (9.5%) may be due to more significant cognitive late effect prohibiting participation. It is important to highlight that this cohort largely represents a group of AYA currently engaged in pediatric survivorship care and therefore may not be representative of all AYA survivors. Of note, self-reported unemployment for AYA was high in this sample (15%); however, this is confounded by the high percentage of students as well as the COVID-19 pandemic. There were significant site differences across both clinical and demographic factors that will be important to consider in future cohort analyses. Demographic differences were expected and was the impetus for recruiting from 3 different geographical regions of the U.S. (Northeast, Midwest and West). Clinical treatment-related and late effects differences across sites were not anticipated but may relate to differing referral and/or transition practices, EHR reporting standards, or catchment areas^64^ which will be important to consider when analyzing and interpreting results.

### FUTURE PLANS

To our knowledge, the AYA TEAMS study is the first prospective cohort of AYA-only off treatment survivors of childhood cancer in the U.S, recruited during the critical developmental period of emerging adulthood and prior to the transition to adult-oriented care. This large cohort is essential for the final validation of the comprehensive Transition Readiness Inventory and to elucidate longitudinal and transactional patterns of, and risk for, poor self-management and disengagement in LTFU. Results from this cohort study will aid in designing tailored and targeted future interventions to enhance AYA self-management, including promotion of sustained engagement in cancer survivorship care.

## Data Availability

All data produced in the present study are available upon reasonable request to the authors.

## Collaboration

The AYA TEAMS database is not open-access due to ethical and data protection constraints. De-identifiable data from this study are available upon reasonable request. Potential collaborators can submit proposals by contacting the principal investigator, Dr. Lisa Schwartz.

## Data sharing statement

De-identifiable data are available upon reasonable request.

## Funding Statement

This work was supported by the National Institute of Nursing Research grant number R01NR017429.

## Acknowledgements

We would like to thank all of the research study staff who assisted with recruitment and data collection: Jordan Wood, Jazmin Trejo, Julia Mahoney, Caitlin Brammer, Courtney Gibson, and all of the AYA and caregivers who participated in this study.

## Authors’ contributions

SKD: data curation, formal analysis, investigation, project administration, supervision, writing - original draft

HF, LG, KN, NR, VF, MJ, BO, AW: investigation, writing - review and editing

KVP, SD, KW: Data curation, investigation, writing - review and editing

DS: Conceptualization, Investigation, methodology, writing - review and editing

JPG, LPB, JAD, KCB, WH: Conceptualization, writing - review and editing

YL, CAT: Conceptualization, methodology, writing - review and editing

DRF, AP: Conceptualization, Funding acquisition, project administration, supervision, writing - review and editing

LAS: Conceptualization, Funding acquisition, Investigation, project administration, supervision, methodology, writing - original draft

## Competing interests statement

The authors have no competing interests to declare.

## Notes

### Competing Interest Statement

The authors have declared no competing interest.

### Author Declarations

The Children's Hospital of Philadelphia IRB gave ethical approval for this work.

